# Healthy lifestyles, plasma proteomic profile and long-term risk of dementia

**DOI:** 10.1101/2024.12.01.24317800

**Authors:** Jie Shen, Minyu Wu, Yuhui Huang, Minqin Yan, Yang Yang, Xin Xu, Geng Zong, Jie Yang, Xue Li, Hui Chen, Changzheng Yuan

## Abstract

Considering the irreversible nature of dementia, lifestyle modification has been recognized as potentially effective preventive strategies. However, whether proteomic signatures of individuals mediate or modifiy the associations between healthy lifestyles and cognitive health remain unclear. Herein, we conducted proteome-wide association analyses and identified 12 plasma proteomic markers that were associated with both overall healthy lifestyle score and incident dementia (Bonferroni corrected *P*-values <0.05) among 53,014 individuals aged 55 years and older in the UK Biobank. Aggregately, they mediated 17.6% (95% CI, 10.0%, 29.6%) of the lifestyle-dementia association. The associations between overall healthy lifestyle score and two key proteinomic markers (GDF15 and IGFBP3) were further replicated in the Framingham Heart Study Offspring cohort. In the proteome-wide interaction analysis, the lifestyle-dementia association significanly differed by certain proteomic features. Using a machine-learning approach, we developed and tested 8 lifestyle protein stratification scores (LPSSs) to identify pariticipants showing stronger associations between overall or individual healthy lifestyles and dementia. In particular, the LPSS for overall healthy lifestyle (consisting of 25 proteins) identified a population with a substantially stronger lifestyle-dementia association (Harzard Ratio: 0.38, 95%CI: 0.32-0.45) compared with overall population (0.67, 0.60-0.75). Individuals with higher LPSS thus may exibit stronger responses to healthy lifestyle interventions, leading to substantially improved proportions of potentially preventable dementia cases. For example, the population attributable fraction of unhealthy diet increased from 1.63% in the overall population to 29.15% in the targeted population with higher LPSS for healthy diet. Our findings highlighted the key proteomic biomarkers that may serve as intervention intermediate outcomes and inform targeted lifestyle interventions for primary prevention of dementia.

## Introduction

Dementia represents a growing global health challenge and greatly impact the functional well-being of older individuals.^1^ As the global population continues to grow and age, the number of people living with dementia is projected to increase from 57.4 million in 2019 to an alarming 152.8 million by 2050.^2^ This surge highlights the urgent need for effective preventive strategies, particularly given the irreversible nature of dementia and the current lack of curative pharmacotherapeutic treatments.^3^ As such, public health efforts have increasingly focused on scaling up interventions through multi-domain lifestyle intervention as a primary preventive measure.^4^ A healthy lifestyle, which encompasses multiple factors such as regular physical activity^5^, a healthy diet^6^, non-smoking^7^, and moderate alcohol consumption^8^, has been consistently associated with a reduced risk of cognitive decline and dementia, irrespective of genetic predisposition of Alzheimer’s disease (AD).^9^ Moreover, an overall healthy lifestyle was associated with a lower risk of AD dementia^10^ and longer life expectancy without AD dementia.^11^ These benefits are largely attributed to cognitive reserve that maintains cognitive abilities independently of common AD pathology.^12^

Despite these well-established associations between healthy lifestyles and cognitive health, whether proteomic signatures mediate or modifiy the associations remain largely unexplored, which is critical for elucidating potential pathways and developing targeted interventions. Specifically, the multiple molecular level manifestations^13^ of protein function or expression in dementia pathogenesis suggest that proteins may play a key role in mediating these lifestyle-dementia associations. Recent advancements in high-dimensional proteomic technologies enable simultaneous measurement of multiple proteins and offer new opportunities for pathway exploration with plasma,^14,15^ cerebrospinal fluid (CSF),^16^ or brain tissue samples.^17^ For example, a longitudinal study leveraging plasma proteomics identified the key roles of proteostasis, immunity, synaptic function, and extracellular matrix organization in dementia development.^15^ However, few studies have investigated the proteomic pathways linking lifestyle factors to dementia. Identifying specific plasma proteins involved in these pathways could reveal potential indicators and intervention targets, potentially enhancing the ability to may predict the efficacy of lifestyle interventions for dementia prevention.

Existing lifestyle intervention studies often yield inconsistent findings, suggesting that the effect may differ by specific protein features of individuals. The success of the two-year Finnish Geriatric Intervention Study to Prevent Cognitive Impairment and Disability (FINGER) proved the potential efficacy of the multidomain intervention in maintaining or improving cognitive function.^18^ However, the Japan-Multimodal Intervention Trial for the Prevention of Dementia (J-MINT), part of the global FINGER network, reported no significant differences in cognitive outcomes between the intervention and control groups. Nevertheless, participants with elevated glial fibrillary acidic protein (GFAP) levels experienced greater and significant benefits from the intervention.^19^ These mixed results underscore the heterogeneity in individual responses to lifestyle interventions, suggesting that a one-size-fits-all approach may be insufficient for dementia prevention at the population level. Tailoring interventions to high-responsiveness populations could enhance the effectiveness of dementia prevention programs, thereby maximizing their preventive potentials.

In this study, we conducted comprehensive proteome-wide association analyses to identify key plasma proteins that mediate the association between an overall healthy lifestyle and the risk of all-cause dementia. Utilizing data from the UK Biobank, we aimed to elucidate the protein mediators of lifestyle-dementia associations that may serve as intervention intermediate outcomes. Further, we aimed to develop and test a set of lifestyle protein stratification scores (LPSSs) that distinguish targeted populations with high responsiveness of overall and individual lifestyle interventions. By identifying critical proteins and constructing stratification scores, our work aim to advance precision prevention of dementia, and assist targeted interventions to preserve cognition in aging populations.

## Results

### Schematic overview and characteristics of study participants

**Figure 1** shows an overview of the study design and analytical approaches. The two major aims are 1) to identify plasma proteins that mediate the association of an overall healthy lifestyle score and its components with incident dementia and 2) to develop protein-based stratification scores to identify targeted populations with stronger associations of certain lifestyles with incident dementia. We considered seven previously documented healthy lifestyles, including no current smoking, regular physical activity, healthy diet, moderate alcohol consumption, healthy sleeping duration, low sedentary behavior, and adequate social activities, and constructed a healthy lifestyle score as described in **Methods** and **Supplementary Table 1.** The ascertainment of incident all-cause dementia was described in **Methods** and **Supplementary Table 2**.

**Figure 1.**
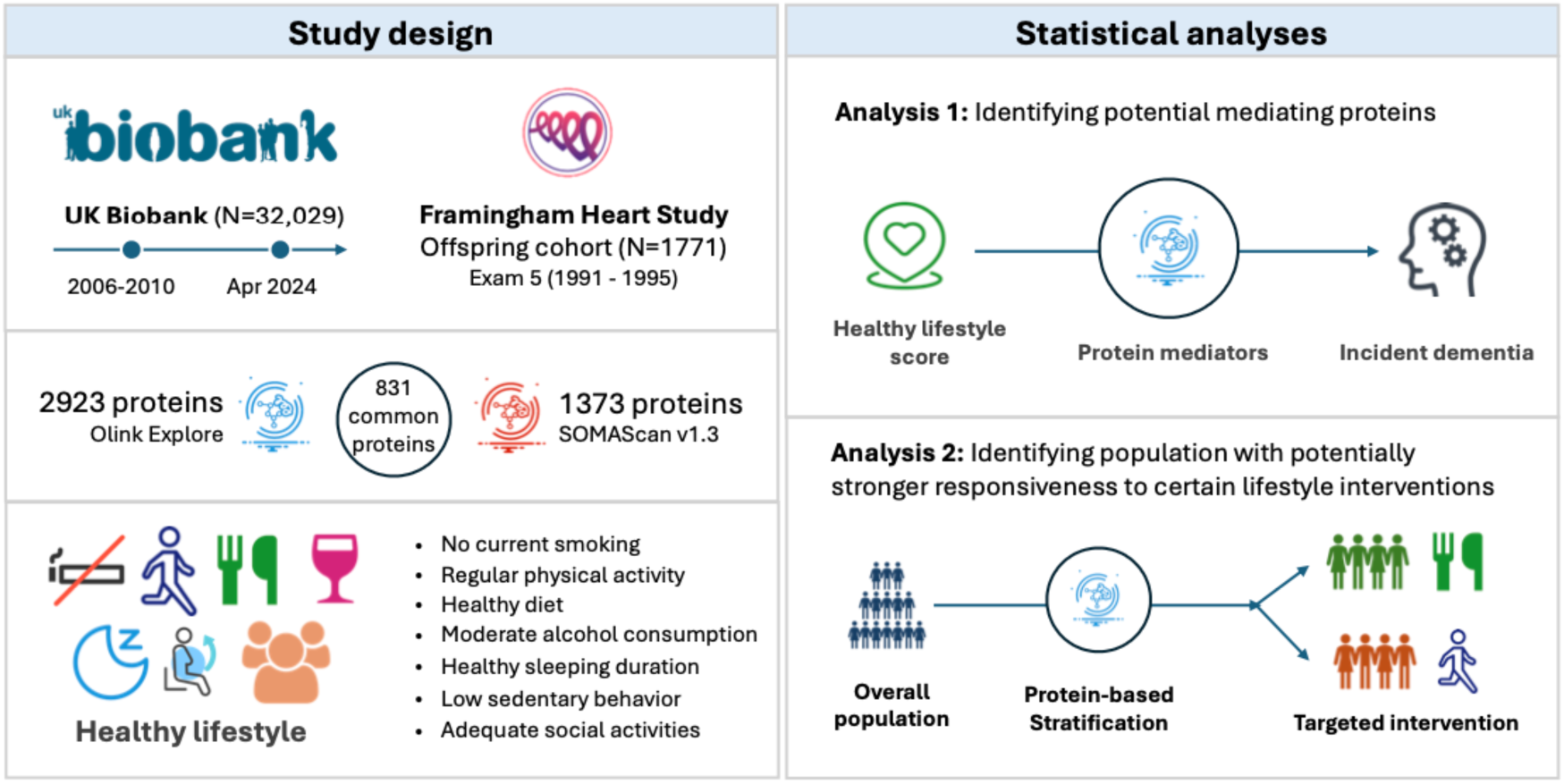
Schematic Overview of Study Design Our analyses were based on 32,029 UK Biobank participants aged 55 years or older in the Pharma Proteomics Project (PPP). Adherence to an overall healthy lifestyle was measured by a weighted composite healthy lifestyle score. Baseline characteristics of the study participants according to healthy lifestyle score quintiles are summarized in ***Table 1***. Participants’ mean age at recruitment was 62.2 years (standard deviation, 4.2), the proportion of females was 53.7%, and most participants (96.2%) self-identified as White ethnicity. Participants in the top quintile of the healthy lifestyle score were more likely to have a college or university degree (39.3%) compared to the bottom quintile (18.9%). Participants with a healthier lifestyle had lower body mass index (BMI) and prevalence of chronic conditions such as diabetes, hypertension, cardiovascular disease, and dyslipidaemia.

**Table 1.**
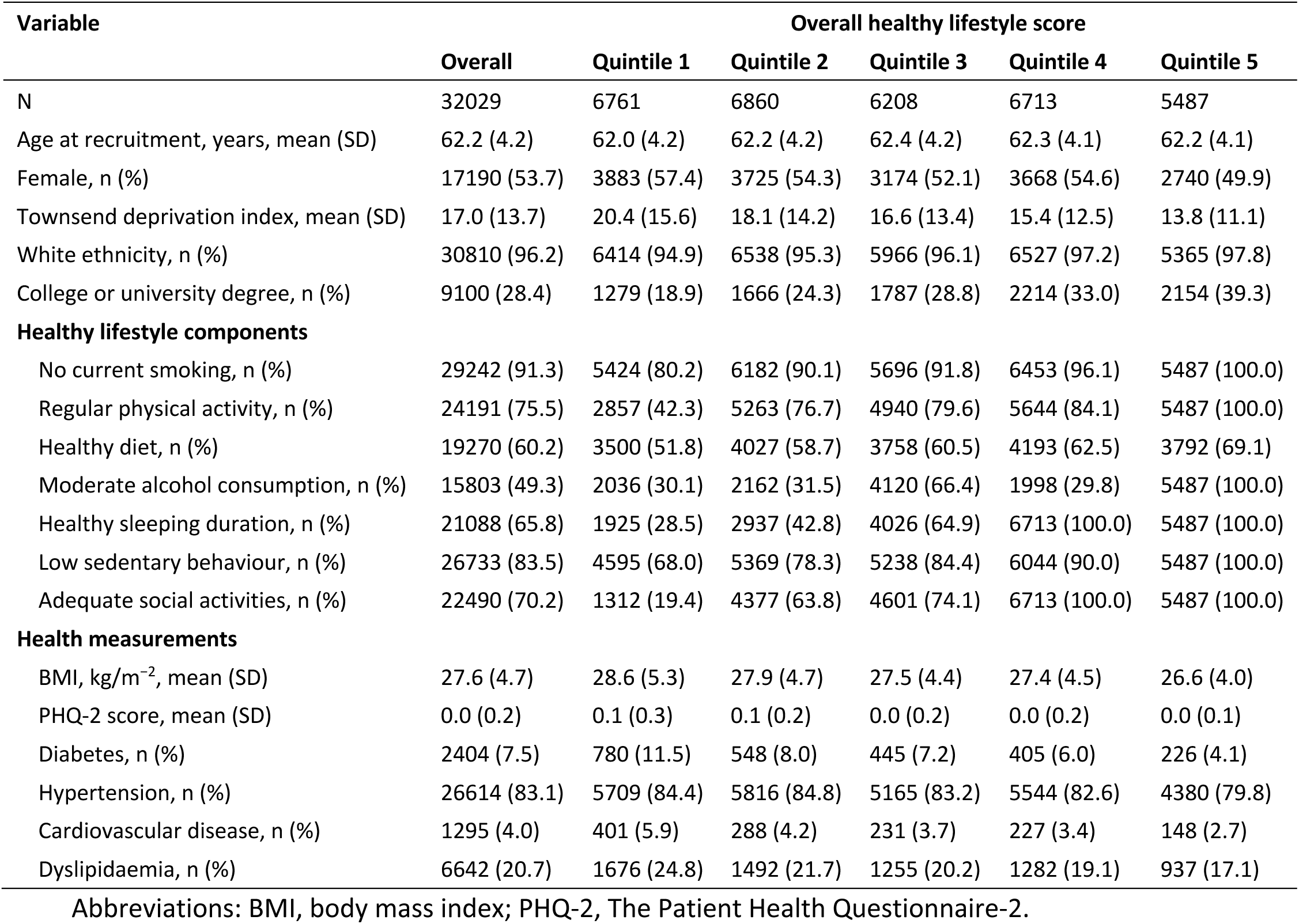
Baseline characteristics of the study participants according to healthy lifestyle score quintiles in UK Biobank.

| Variable | Overall healthy lifestyle score |  |  |  |  |  |
| --- | --- | --- | --- | --- | --- | --- |
|  | Overall | Quintile 1 | Quintile 2 | Quintile 3 | Quintile 4 | Quintile 5 |
| N | 32029 | 6761 | 6860 | 6208 | 6713 | 5487 |
| Age at recruitment, years, mean (SD) | 62.2 (4.2) | 62.0 (4.2) | 62.2 (4.2) | 62.4 (4.2) | 62.3 (4.1) | 62.2 (4.1) |
| Female, n (%) | 17190 (53.7) | 3883 (57.4) | 3725 (54.3) | 3174 (52.1) | 3668 (54.6) | 2740 (49.9) |
| Townsend deprivation index, mean (SD) | 17.0 (13.7) | 20.4 (15.6) | 18.1 (14.2) | 16.6 (13.4) | 15.4 (12.5) | 13.8 (11.1) |
| White ethnicity, n (%) | 30810 (96.2) | 6414 (94.9) | 6538 (95.3) | 5966 (96.1) | 6527 (97.2) | 5365 (97.8) |
| College or university degree, n (%) | 9100 (28.4) | 1279 (18.9) | 1666 (24.3) | 1787 (28.8) | 2214 (33.0) | 2154 (39.3) |
| **Healthy lifestyle components** |  |  |  |  |  |  |
| No current smoking, n (%) | 29242 (91.3) | 5424 (80.2) | 6182 (90.1) | 5696 (91.8) | 6453 (96.1) | 5487 (100.0) |
| Regular physical activity, n (%) | 24191 (75.5) | 2857 (42.3) | 5263 (76.7) | 4940 (79.6) | 5644 (84.1) | 5487 (100.0) |
| Healthy diet, n (%) | 19270 (60.2) | 3500 (51.8) | 4027 (58.7) | 3758 (60.5) | 4193 (62.5) | 3792 (69.1) |
| Moderate alcohol consumption, n (%) | 15803 (49.3) | 2036 (30.1) | 2162 (31.5) | 4120 (66.4) | 1998 (29.8) | 5487 (100.0) |
| Healthy sleeping duration, n (%) | 21088 (65.8) | 1925 (28.5) | 2937 (42.8) | 4026 (64.9) | 6713 (100.0) | 5487 (100.0) |
| Low sedentary behaviour, n (%) | 26733 (83.5) | 4595 (68.0) | 5369 (78.3) | 5238 (84.4) | 6044 (90.0) | 5487 (100.0) |
| Adequate social activities, n (%) | 22490 (70.2) | 1312 (19.4) | 4377 (63.8) | 4601 (74.1) | 6713 (100.0) | 5487 (100.0) |
| **Health measurements** |  |  |  |  |  |  |
| BMI, kg/m<sup>-2</sup>, mean (SD) | 27.6 (4.7) | 28.6 (5.3) | 27.9 (4.7) | 27.5 (4.4) | 27.4 (4.5) | 26.6 (4.0) |
| PHQ-2 score, mean (SD) | 0.0 (0.2) | 0.1 (0.3) | 0.1 (0.2) | 0.0 (0.2) | 0.0 (0.2) | 0.0 (0.1) |
| Diabetes, n (%) | 2404 (7.5) | 780 (11.5) | 548 (8.0) | 445 (7.2) | 405 (6.0) | 226 (4.1) |
| Hypertension, n (%) | 26614 (83.1) | 5709 (84.4) | 5816 (84.8) | 5165 (83.2) | 5544 (82.6) | 4380 (79.8) |
| Cardiovascular disease, n (%) | 1295 (4.0) | 401 (5.9) | 288 (4.2) | 231 (3.7) | 227 (3.4) | 148 (2.7) |
| Dyslipidaemia, n (%) | 6642 (20.7) | 1676 (24.8) | 1492 (21.7) | 1255 (20.2) | 1282 (19.1) | 937 (17.1) |

Abbreviations: BMI, body mass index; PHQ-2, The Patient Health Questionnaire-2.

### Healthy lifestyles associated with lower all-cause dementia risk

During 419,745 person-years (mean follow-up = 13.1 years), we documented 1,296 incident all-cause dementia cases (participant characteristics described in **Supplementary Table 3**). An overall healthy lifestyle was associated with a lower risk of all-cause dementia (P-trend <0.001, **Figure 2**a) in a dose-response manner (**Figure 2**b). Participants in the highest quintile of overall healthy lifestyle score had a 49% lower dementia risk (Harzard Ratio [HR] = 0.51, 95% CI: 0.42-0.62) compared to those in the lowest quintile. Each quintile increment in the overall healthy lifestyle score was related to a 16% reduction in risk of dementia (HR = 0.84, 95% CI: 0.80-0.87).

**Figure 2.**
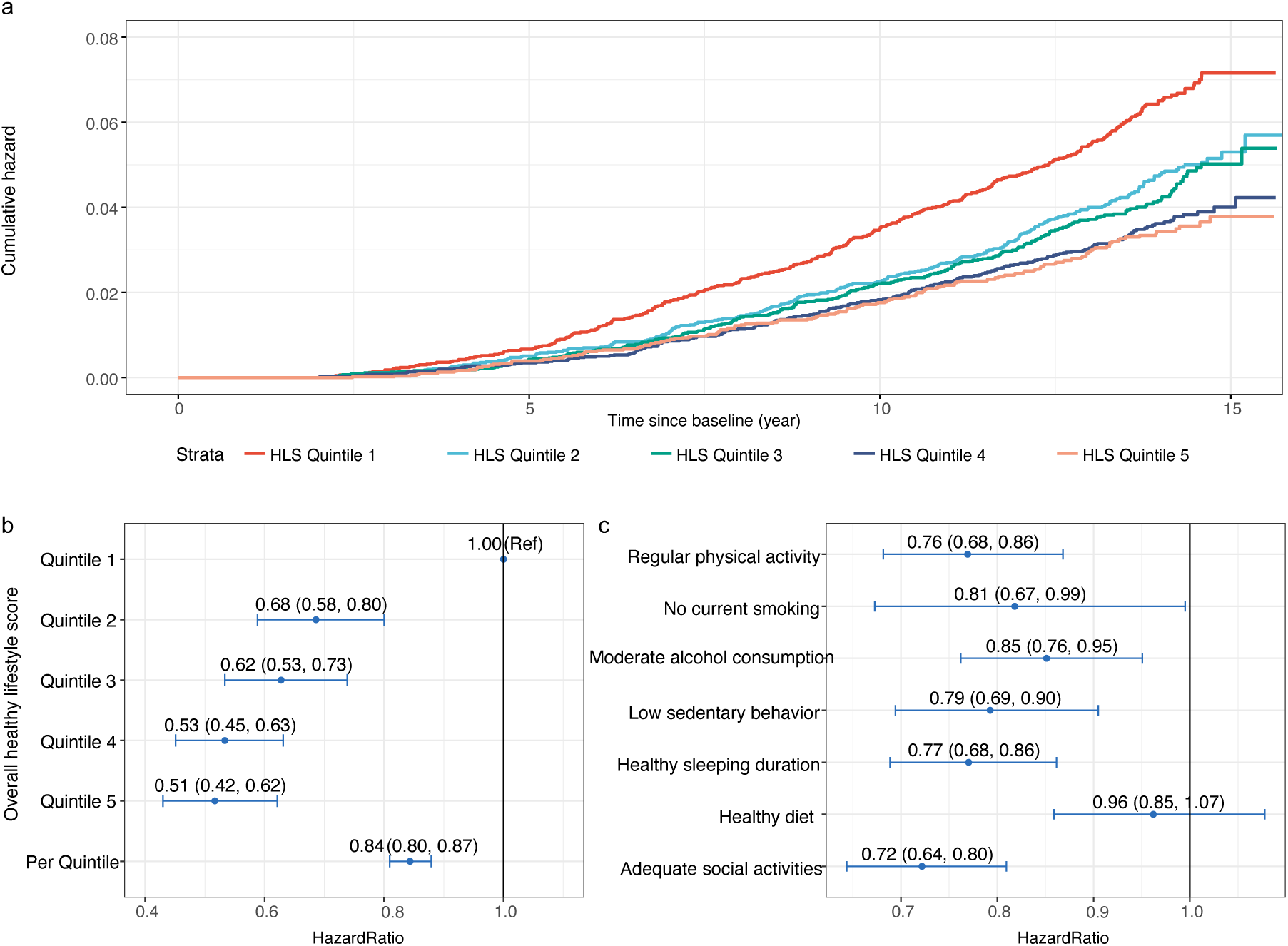
Associations of healthy lifestyle score and individual lifestyle components with incident dementia a. Cumulative hazard of incident dementia according to healthy lifestyle score in quintiles; b. Association of the health lifestyle score with incident dementia; c. Association of individual healthy lifestyle factors with incident dementia; Healthy lifestyle score was categorized into quintiles and the lowest group (indicating participants with the worst lifestyle) was set as the reference group. Hazard ratios and 95% confidence intervals were calculated from the Cox proportional hazard models adjusted for age at recruitment, sex, ethnicity, Townsend deprivation index, and highest education level.

**Figure 2**c presents the independent associations of individual healthy lifestyle factors with the risk of dementia. Regular physical activity, no current smoking, moderate alcohol consumption, low sedentary behaviour, healthy sleeping duration, and adequate social activities were independently significantly associated with a lower risk of dementia, with HRs ranging from 0.72 (95% CI, 0.64, 0.80) for adequate social activities to 0.85 (95% CI, 0.76, 0.95) for moderate alcohol consumption. These findings suggest that while multiple lifestyle components contribute to lowering dementia risk, their relative impacts vary. In addition, healthy diet was not independently associated with risk of dementia in the multivariable adjusted model (HR = 0.96, 95% CI: 0.85-1.07).

### Plasma proteins link healthy lifestyles to future risk of all-cause dementia

The UK Biobank Pharma Proteomics Project (PPP) quantified 2,923 plasma proteins using the Olink Explore panels, of which 2,911 proteins with missing rates less than 20% were included in our analysis. We assessed their associations with the healthy lifestyle score using linear regression models (**Supplementary Table 4**). A total of 406 plasma proteins were significantly associated with the overall healthy lifestyle score (Bonferroni-corrected P-values <0.05, **Figure 3**a). Using Cox proportional hazard models, we further identified 23 plasma proteins that were significantly associated with future risk of incident all-cause dementia (Bonferroni-corrected P-values <0.05, **Figure 3**b). The strongest associations were detected for the GFAP, neurofilament light polypeptide (NEFL), and growth/differentiation factor 15 (GDF15), which have been previously documented.^15,16,20^

**Figure 3.**
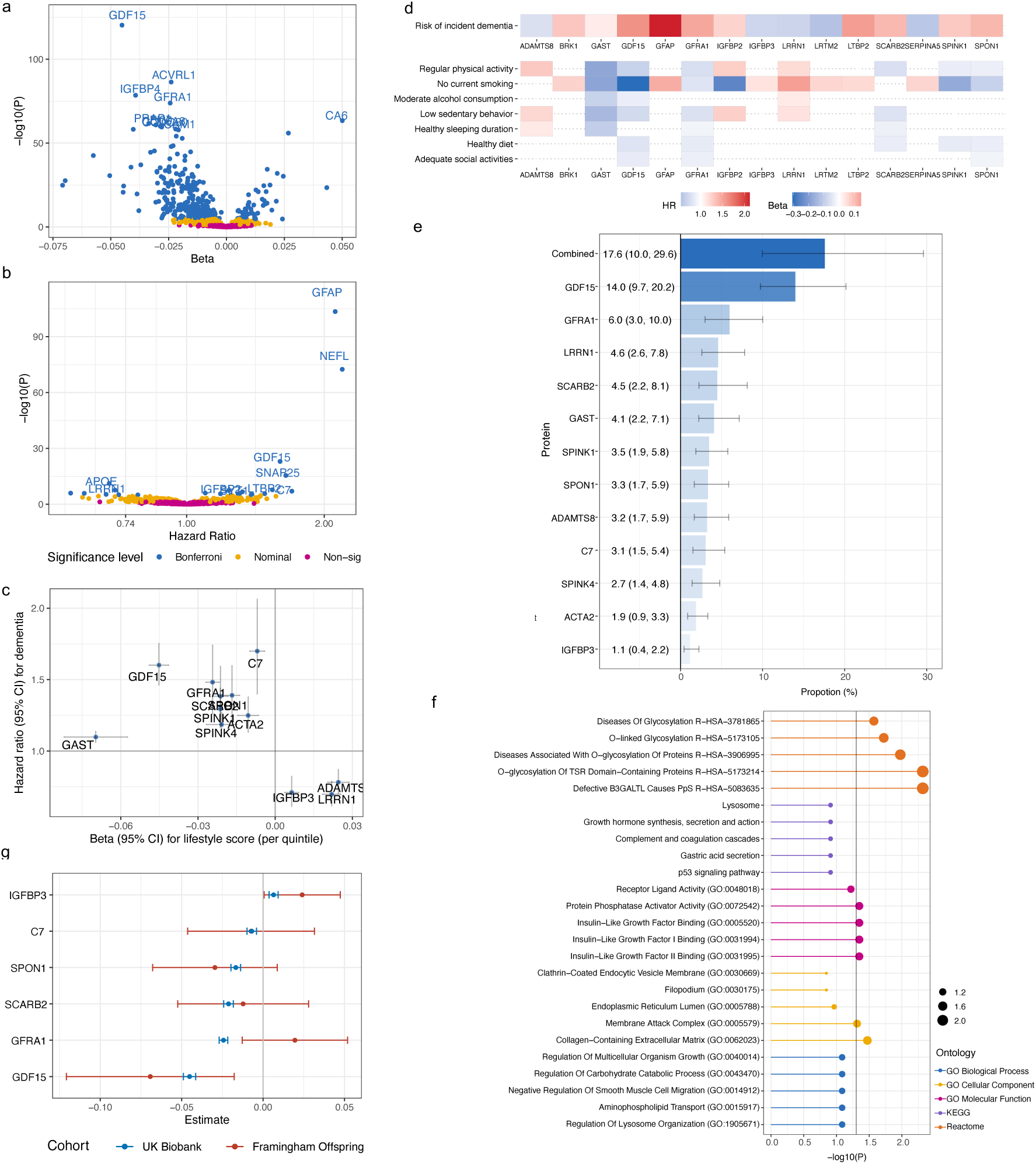
Identification and validation of mediating proteins in the association between healthy lifestyle score and incident dementia and enrichment analysis a. Volcano plot showing the beta coefficients (x axis) and −log10(P value) (y axis) for the associations of healthy lifestyle score with 2,911 plasma proteins calculated from linear regression models; b. Volcano plot showing the hazard ratios (x axis) and −log10(P value) (y axis) for the associations of 2,911 plasma proteins with incident dementia calculated from Cox proportional hazard models; c. Scatter plot showing the beta coefficients (x axis) and hazard ratios (y axis) of 12 plasma proteins that are associated with both healthy lifestyle score and incident dementia (Spearman correlation=-0.47, indicating that proteins associated with healthier lifestyle were associated with lower dementia risk). d. Plasma proteins associated with healthy lifestyle component and incident dementia. e. Mediation proportion (95% confidence intervals) of the 12 plasma proteins that are associated with both healthy lifestyle score and incident dementia. All analyses were adjusted for age at recruitment, sex, ethnicity, Townsend deprivation index, and highest education level. Bonferroni-corrected P < 0.05 indeicated statistical significance for all tests. f. Enrichment for Gene Ontology (GO), Kyoto Encyclopedia of Genes and Genomes (KEGG) and Reactome pathways from the 12 plasma proteins that are associated with both healthy lifestyle score and incident dementia. Point sizes indicated -log_10_(FDR corrected P-value), and the vertical line indicated the significance threshold for FDR correction. g. External validation of the identified proteins in the FOS, where 6 out of the 12 proteins were measured using SOMAScan v1.3. All analyses were adjusted for age at recruitment, sex, ethnicity, Townsend deprivation index (or household income), and highest education level. Bonferroni-corrected P < 0.05 indicated statistical significance for all tests.

Combining the large-scale proteome-wide association analyses of overall healthy lifestyle and dementia risk, we identified 12 proteins significantly associated with both the overall healthy lifestyle score and dementia risk (**Figure 3**c). Among them, ADAMTS8, LRRN1, IGFBP3 were associated with higher overall healthy lifestyle score and lower dementia risk, and GAST, GDF15, GFRA1, SPINK1, SCARB2, SPINK4, SPON1, ACTA2 and C7 were associated with a lower overall healthy lifestyle score and higher dementia risk. We further identified proteins that were linked to individual healthy lifestyle components, detailed in **Supplementary Table 5** and **Figure 3**d. In addition to the 12 proteins, BRK1, GFAP, IGFBP2, LRTM2, LTBP2, and SERPINA5 were also associated with certain individual lifestyle factors. Notably, all seven healthy lifestyle factors were significantly associated with lower level of GDF15.

Furthermore, we quantified the mediation proportion of the 12 identified proteins in the lifestyle-dementia association in the UK Biobank (**Figure 3**e), with mediation proportions ranging from 1.1% (IGFBP3) to 14.0% (GDF15). Collectively, they mediated 17.6% of the association between the overall lifestyle score and dementia risk. In addition, we explored the enriched pathways through which healthy lifestyle may influence future dementia risk (**Figure 3**f). The Reactome pathways of abnormalities in protein glycosylation processes, addition of sugars to proteins at specific sites (O-linked), and faulty O-glycosylation emphasized the importance of protein glycosylation in lifestyle and dementia, which were further supported by the key roles of insulin-life growth factor-related molecular-level pathways identified by GO. At cellular level, membrane-attack complex and collagen-containing extracellular matrix pathways were involved. The KEGG pathways of immune regulation and inflammation and p53 signalling pathway also suggested a potential role of apoptosis. The protein–protein interaction network of the 12 proteins constructed based on the STRING database is shown in **Supplementary Figure 2**. The network is featured by a central role of the GDF15 (connected to SPON1, GFRA1, and IGFBP3), along with the SPINK1- SPIN4 and SCARB2-ADAMTS8 connections.

We externally replicated the analysis for 6 of them which were also profiled in the FOS (N = 1771; participant characteristics described in **Supplementary Table 6**) using the aptamer-based SOMAScan® v1.3 proteomics platform. The overall healthy lifestyle score was significantly associated with higher IGFBP3 level and lower GDF15 level (**Figure 3**g). While other proteins did not demonstrate significant associations, most point estimates indicated similar directions to the results from UK Biobank.

### Protein-based scores stratify populations according to strengths of lifestyle-dementia associations

In the proteome-wide interaction analysis, we identified key proteins that significantly modified the associations of the overall healthy lifestyle score and its components with incident dementia (**Supplementary Table 7**). We adopted a conservative approach to detect potential interactions between proteins and lifestyle factors by defining statistical significance at a *P*-interaction <0.10.^21–23^

From the 333 proteins showing nominal significance in the interaction with healthy lifestyle score, we developed a Lifestyle Protein Stratification Score (LPSS) for the overall healthy lifestyle score (LPSS-Overall, consisting of 25 proteins) that stratified the associations of the overall healthy lifestyle score with future risk of dementia, using a forward stepwise approach in a random training set consisting of 50% of the population. This strategy allowed us to identify populations with potentially varying responsiveness to an overall healthy lifestyle (**Figure 4**a and **Figure 4**b). Specifically, participants with a higher LPSS-Overall had a stronger lifestyle-dementia associations. The HR for high v. low (defined by median) overall healthy lifestyle score strengthened from 0.67 (0.60-0.75) in the overall population to 0.38 (0.32-0.45) in the targeted population with a high LPSS-Overall (*P*-interaction = 3.3e-20). Therefore, they may benefit more from the overall ifestyle intervention. This was further confirmed in a separate testing set (HR strengthed from 0.69, 0.59-0.81 in the overall population to 0.56, 0.45-0.71 in the targeted population (*P*-interaction = 0.01, **Figure 4**c).

**Figure 4.**
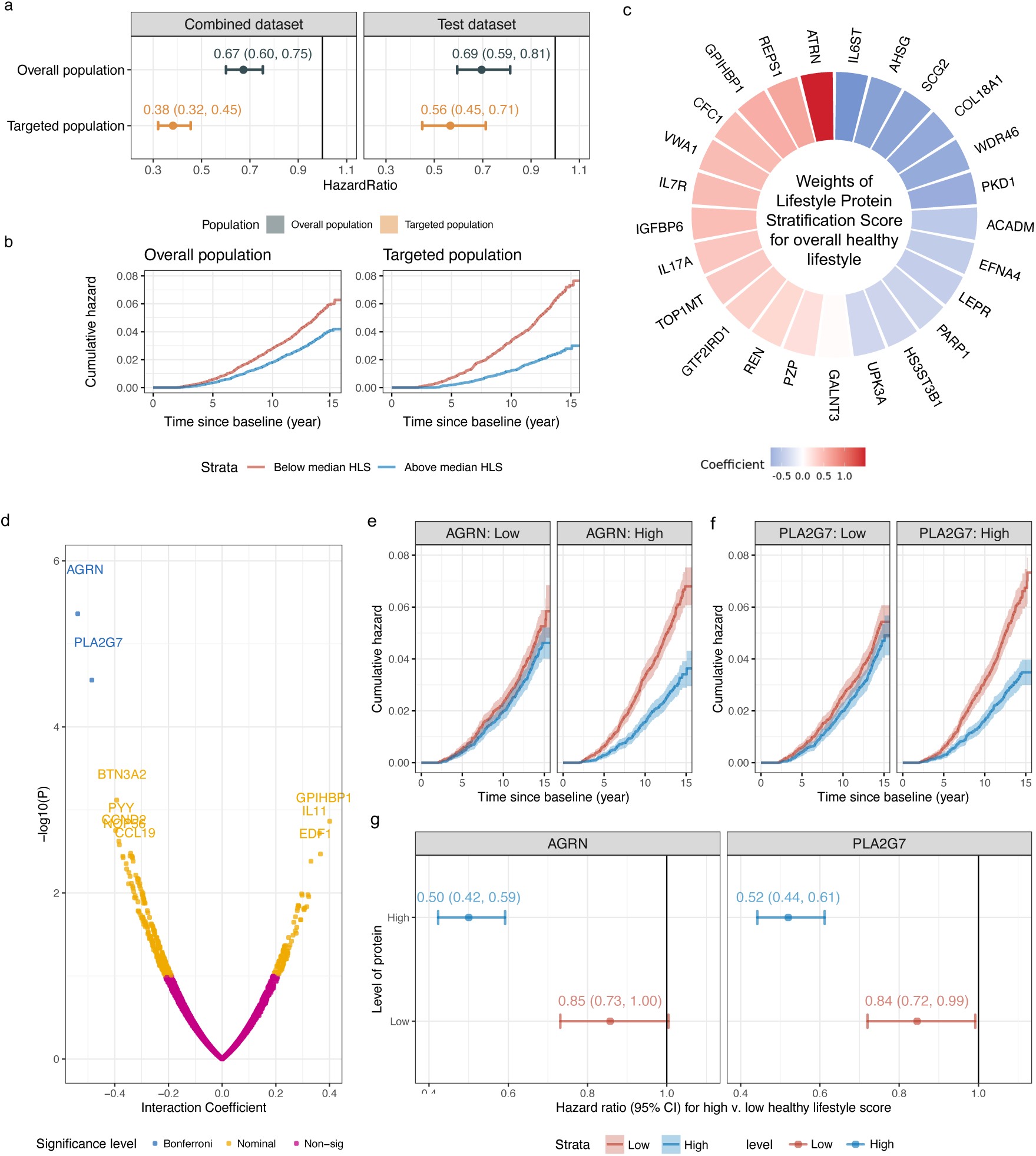
Plasma proteins and lifestyle protein stratification score modifying the association between healthy lifestyle score and incident dementia **a**. Association of healthy lifestyle score with risk of incident all-cause dementia in the overall population (blue) and targeted population (yellow), with the *P* values for interaction for LPSS-Overall being 3.3e-20 in the combined set and 0.01 in the testing set; **b**. Cumulative hazard of incident dementia according to healthy lifestyle score in the overall and targeted population; **c**. The weights of LPSS-Overall derived in traning set. All analyses were adjusted for age at recruitment, sex, ethnicity, Townsend deprivation index, and highest education level; **d**. Volcano plot showing the beta coefficients (x axis) and −log10(P value) (y axis) for the interaction term crossing healthy lifestyle score with 2,911 plasma proteins from Cox proportional hazard models; **e**. Cumulative hazard of incident dementia according to the healthy lifestyle score by plasma AGRN levels; **f**. Cumulative hazard of incident dementia according to the overall healthy lifestyle score by plasma PLA2G7 levels; g. The overall healthy lifestyle score and plasma proteins were categorized according to median values. All analyses were adjusted for age at recruitment, sex, ethnicity, Townsend deprivation index, and highest education level. Bonferroni-corrected P-interaction <0.10 indicated statistical significance for healthy lifestyle score × protein interaction.

Among the 333 proteins that showing nominal significance in the interaction with healthy lifestyle score, agrin (AGRN, *P*-value=4.4e-06) and platelet-activating factor acetylhydrolase (PLA2G7, *P*-value=2.7e-05) further passed Bonferroni correction (Bonferroni-corrected *P*- interactions = 0.012 and 0.079, respectively). Participants with higher AGRN and PLA2G7 levels had significantly stronger overall lifestyle-dementia associations (HR comparing high v. low healthy lifestyle score = 0.50, 95% CI: 0.42-1.59 in high AGRN and 0.52, 0.44-0.61 in high PLA2G7) than those with lower levels (0.85, 0.73-1.00 in low AGRN and 0.84, 0.72-0.99 in low PLA2G7) (**Figure 4**e-g). These findings suggest that certain proteins may help identify populations who may have higher response to lifestyle interventions, thus improving the potential effectiveness in real-world practices.

Similarly, we constructed 7 separate LPSSs for individual healthy lifestyles to stratify individuals with stronger associations of specific healthy lifestyle factors (**Figure 5**a and **Figure 5**b-h). Each LPSS for individual healthy lifestyle consisted of 17-34 proteins. Among them, the LPSS for low sedentary behavior (LPSS-Sedentary), using 33 proteins, exhibited exceptional densitfication performance (*P*-interaction = 4.0e-25 for combined set; *P*- interaction = 5.7e-05 for testing set). The decreased risk of dementia that associated with low sedentary behavior was stronger in the targeted population with higher LPSS-Sedentary score (HR _combined_ = 0.42, 95% CI: 0.35-0.49; HR _testing_ = 0.60, 0.46-0.77) than those in the overall population (HR _combined set_ = 0.79, 0.69-0.90; HR _testing_ = 0.87, 0.72-1.05). Interestingly, the LPSS for healthy diet (LPSS-Diet), based on 28 proteins, identified populations with a significant and strong association between a healthy diet and all-cause dementia (HR _combined_ = 0.63, 0.54-0.74, *P*-interaction=1.8e-15; HR _testing_ = 0.76, 0.61-0.95, *P*-interaction=0.01), despite non-significant association observed in the overall population. The weights of the LPSSs for individual healthy lifestyles were shown in **Figure 5**j and **Supplementary Table 8.** The population attributable fraction (PAF) for dementia due to each unhealthy lifestyle factor was higher in the targeted population with higher LPSSs (ranging from 12.10% to 29.15%) compared to that in the overall population (ranging from 1.63% to 10.39%) (**Figure 5**i). Namely, the LPSSs for overall healthy lifestyle score and individual healthy lifestyles improved the proportions of potentially reduceable dementia cases among the targeted populations. For example, the PAF of unhealthy diet increeased from 1.63% (non-significant) in the overall population to 29.15% in the targeted population, which suggested that if all adopting a healthy diet, the targeted population may have ∼30% fewer dementia cases. The interactions of proteins and each individual healthy lifestyle factors was shown in **Supplementary Table 9.**

**Figure 5.**
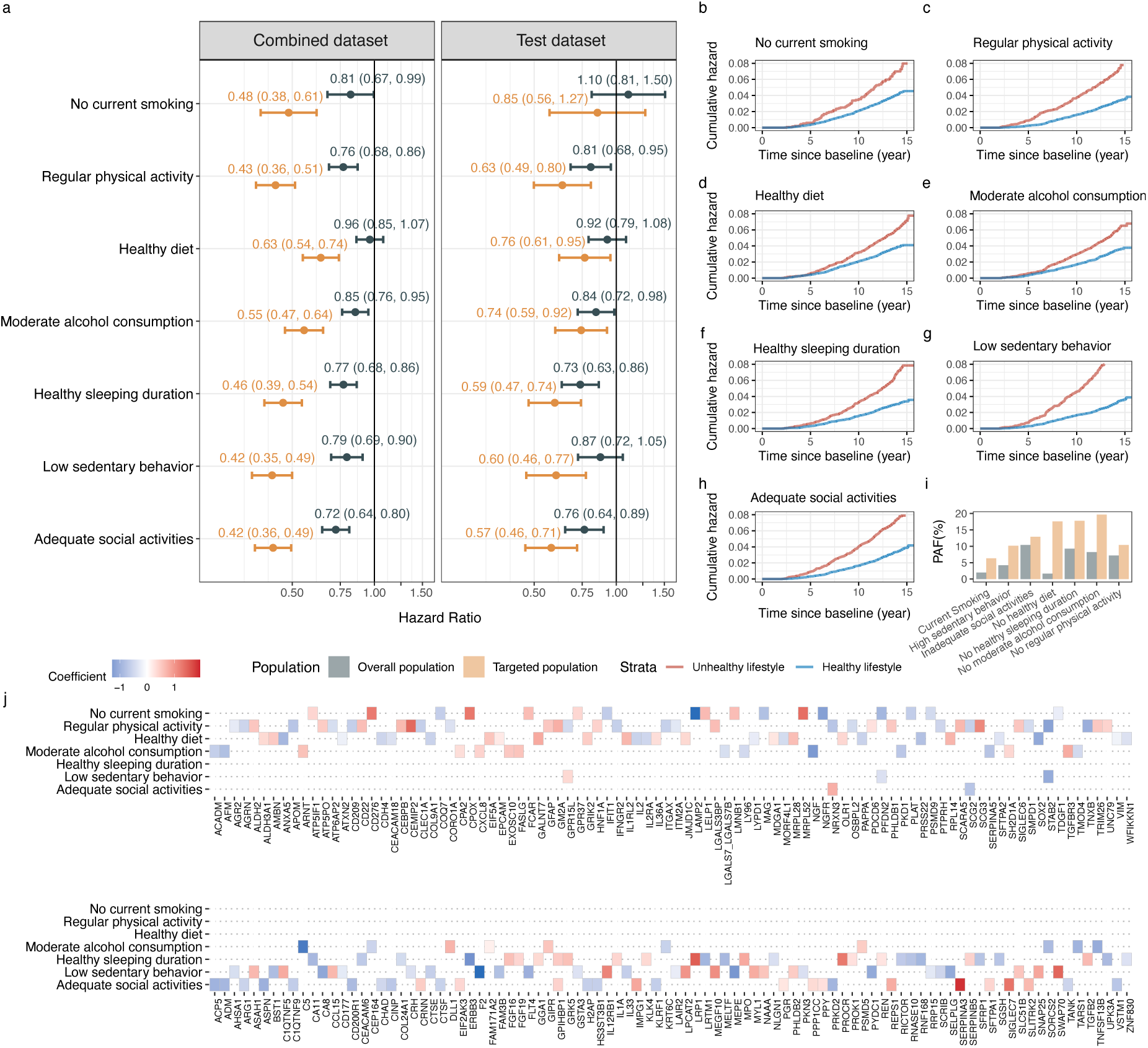
Lifestyle Protein Stratification Scores (LPSSs) to prioritize participants with greater responsiveness of individual healthy lifestyle factors on dementia risk **a**. Association of healthy lifestyle factors with risk of incident all-cause dementia in all population (blue) and targeted population (yellow); **b-h**. Cumulative hazard of incident dementia according to healthy lifestyle factors in targeted population, red indicating unhealthy lifestyle and blue indicating healthy lifestyle; **i**. Population attributable fraction (% PAF) for lifestyle risk factors; **j**. The weighting coeffients for separate healthy lifestyle factors. All analyses were adjusted for age at recruitment, sex, ethnicity, Townsend deprivation index, and highest education level. In combined set, *P* values for interaction for Lifestyle × LPSSs were 1.5e-09 for no current smoking, 1.8e-15 for healthy diet, 1.1e-20 for regular physical activity, 2.8e-14 for moderate alcohol consumption, 4.5e-19 for healthy sleeping duration, 4.0e-25 for sedentary behaviour, and 6.7e-12 for adequate social activities. In testing set, *P* values for interaction for PPSSs were 0.08 for no current smoking, 0.01 for healthy diet, 0.01 for regular physical activity, 0.11 for moderate alcohol consumption, 0.01 for healthy sleeping duration, 5.7e-05 for sedentary behaviour, and 3.1e-04 for adequate social activities.

## Discussion

In this study, we conducted large-scale proteomic analyses in a well-characterized cohort and identified several proteomic markers (e.g., GDF15, IGFBP3) that may serve as mediators in the association between healthy lifestyles and lower risk of incident dementia. We also constructed specific LPSSs to identify populations with stronger lifestyle-dementia associations. In particular, the LPSS for overall healthy lifestyle (LPSS-Overall, consisting of 25 proteins) identified a subpopulation with substantially pronounced assocations between overall healthy lifestyle and lower risk of dementia (HR= 0.38, 95%CI: 0.32-0.45) compared to this in the general population (HR= 0.67, 95%CI: 0.60-0.75)). Our findings highlighted the importance of these key proteomic biomarkers that may serve as intervention intermediate outcomes and inform targeted lifestyle interventions for primary prevention of dementia.

The overall healthy lifestyle and specific healthy lifestyle factors have been extensively associated with a lower risk of dementia and age-related cognitive decline. Among 196,383 UK Biobank participants, a higher healthy lifestyle score was related to a 25% lower risk of dementia, regardless of genetic risk.^9^ Similarly, the Chicago Health and Aging Project, which conducted in 2,449 older adults, reported that participants with more healthy lifestyle factors had longer life expectancy and fewer years lived with Alzheimer’s dementia across the lifespan.^11^ In our previous study, non-smoking, adequate physical activity, moderate alcohol consumption, and adequate sleep duration may be independent protective factors against dementia.^24^ In this study, we further confirmed the independent associations for adequate social activities and low sedentary time, which have been identified as emerging lifestyle factors associated with healthy aging.^25,26^ However, the mechanisms linking these factors to brain aging and neurodegeneration remain largely unexplored.

Compared with brain tissues and CSF, plasma proteome is more feasible for large-scale screening in clinical practice. Plasma proteomic assays also enable discovery of molecular-level changes associated with disease development. Previous studies have identified multiple plasma proteins that were linked to prevalent or incident dementia.^20,27^ For example, in a previous proteomic study using 1,463 proteins based on the UK Biobank, GFAP, NEFL, GDF15 and LTBP2 were associated with incident dementia, among which GFAP and NEFL may potentially predict dementia risk 10 years before dementia.^28^ In the Atherosclerosis Risk in Communities with longitudinal plasma proteomic measurements, GDF15 level was associated with dementia risk in a strongly magnitude, especially with an extended follow-up of >15 years.^14,15^ Our findings largely align with these previous studies and showed that 23 out of 2911 plasma proteins were significantly associated with future risk of all-cause dementia, with GFAP, NEFL, and GDF15 showing the prominent associations.

Based on these findings, we conducted large-scale mediation analyses to identify key proteins that mediated the lifestyle-dementia association, which have been utilized in the investigation of metabolomic profiles of certain risk factors and health outcomes. For example, in four large cohorts with 13,056 individuals and 28-year follow-up, the healthy lifestyle score was related to higher levels of 58 metabolites and lower levels of 129 metabolites, which explained 38% of the lifestyle-mortality associations.^29^ In another study, a metabolic signature of 67 metabolites, reflective of adherence to the Mediterranean diet, was associated with cardiovascular disease risk^30^. Similarly, we have previously reported the mediating roles of metabolomic features of the MIND diet in the diet-dementia associations^31^. In a previous study in the UK Biobank, cardiovascular health was also strongly associated with specific dementia-related proteins.^32^ These findings suggest that large-scale omic-based association analyses can provide valuable mechanism insights in the association between risk factors and dementia. In the current study, we identified 12 proteins that were associated with both healthy lifestyle and future risk of dementia: ADAMTS8, LRRN1, IGFBP3 linked to higher healthy lifestyle score and lower dementia risk, and GAST, GDF15, GFRA1, SPINK1, SCARB2, SPINK4, SPON1, ACTA2 and C7 exhibited reverse associations. All of them have been associated with dementia pathogenesis^15,33–36^. Enrichment analyses further indicated the central roles of abnormalities in protein glycosylation processes and apoptosis regulation in the association between lifestyle and dementia. Specifically, GDF15, a member of the transforming growth factor-β, plays essential role in cell multiplication, differentiation, and repair and showed high differential utilities in neurodegenerative diseases.^37^ IGFBP3, which is also externally validated in the current study, has also been linked to reduced oxidative stress and longevity with its neurotrophic properties.^35^

In addition to reflecting the status of specific risk factors as relevant intervention targets, circulating proteins may also assist in identifying specific high-responsiveness populations. For example, in the J-MINT, where multi-domain intervention did not significantly improve cognition in the overall at-risk population as compared with the control strategies, investigators found a significant effect among individuals with a high GFAP level.^19^ Similarly, in the MIND trial that focused on dietary intervention, individuals with higher levels of NEFL, pTau-181 and GFAP responded to the intervention with larger and significant cognitive benefits,^38^ although the overall population did not demonstrate a significant effect.^39^ These findings suggest that overall or specific lifestyle interventions may have considerable heterogeneity across the population, and proteins may be important markers to identify targeted populations and explain this heterogeneity. Therefore, we further constructed a set of LPSSs for overall and individual healthy lifestyles that may help identify high-responsiveness populations, attempting to explore the feasibility of precision intervention. These scores efficiently stratify populations according to their lifestyle-dementia associations, potentially identifying populations that may benefit more from overall or specific lifestyle intervention to promote precise prevention of dementia. While the LPSSs are expected to be further validated and calibrated, they may indicate potentially greater response to healthy lifestyle for dementia.

In addition, we demonstrated that AGRN and PLA2G7 were two key proteins that may modify the association between overall healthy lifestyle intervention and dementia. The AGRN has been reported to promote heart regeneration in adult mice after myocardial infarction through the disassembly of the dystrophin–glycoprotein complex.^40^ Our study further suggests that participants with a higher AGRN level may have a stronger lifestyle-dementia association. PLA2G7 is involved in phospholipid catabolism during inflammatory and oxidative stress responses^41^. A previous study has reported that restricted caloric intake benefited health span and life span in model organisms, in mice, and in healthy humans, through regulations of the PLA2G7.^42^ Our findings suggest that individuals with higher levels of PLA2G7 were more likely to respond favourably to overall lifestyle changes, further supporting the important role of inflammation and oxidative inhibition in brain aging.

However, our findings should be interpreted with caution due to several limitations. Our study relies on observational cohorts, which introduces potential reverse causation and residual confounding that could bias the associations. Therefore, the large-scale association and interaction analyses do not necessarily imply causal linkages, which could only be confirmed by randomized clinical trials. Secondly, the proteins included in the Olink Explore 3072 assay in the UK Biobank covered only ∼3000 proteins, with <30% overalapping proteins the SOMAScan v1.3 panel (∼1300 proteins). As a result, we could not capture and validate all proteins that may explain the lifestyle-dementia associations. Future advancements in the proteomic technology may further improve the performance of our approaches. Finally, given the limited sample size of the FOS, we were only able to verify associations between lifestyle and specific proteins. Future studies are needed to further confirm our other findings. Our study has multiple strengths, including a large sample size, long-term follow-up for dementia, well-characterized populations, and deeply profiled proteomic data that facilitated discovery of multiple proteins. In addition, the LPSSs mark a novelty to improve cost-effectiveness in intervention design, if proven feasible and replicable.

In summary, this study indentified a set of protomic markers (e.g. GDF15 and IGFBP3) that may serve as potential mediators in the associations between healthy lifestyles and dementia Overall and specific lifestyle protein stratification scores were constructed and utilized to identify populations with a stronger lifestyle-dementia association. These findings highlighted the key proteomic biomarkers that may serve as intervention intermediate outcomes and inform targeted and personzlied lifestyle interventions for primary prevention of dementia.

## Data Availability

All data produced in the present study are available upon reasonable request to the authors

## Acknowledgements

We thank all of the participating individuals and staff of the UK Biobank and the Framingham Heart Study who made the study possible. No financial compensation beyond usual salary was received for the provided contributions.

## Author contributions

CY and HC concepted and designed the study and take responsibility for the integrity of the data and the accuracy of the data analysis. Acquisition, analysis, or interpretation of data: HC, JS. Statistical analysis: HC, JS. Drafting of the manuscript: HC, JS, MY. Administrative, technical, or material support: CY, HC. All authors critically revised the manuscript for important intellectual content and read and approved the final manuscript.

## Resource availability

### Lead contact

Further information and requests for resources and reagents should be directed to and will be fulfilled by the lead contact, Changzheng yuan.

## Materials availability

This study did not generate new unique reagents.

## Data and code availability

The main data used in this study were accessed from the UK Biobank (https://biobank.ndph.ox.ac.uk/) and FOS (https://www.framinghamheartstudy.org/fhs-forresearchers/). UKB data are publicly available to bona fide researchers upon application at http://www.ukbiobank.ac.uk/using-the-resource/.

## Methods

### Study Participants

The UK Biobank is a population-based cohort of over 0.5 million residents aged 37-69 years in 2006-2010 (https://biobank.ndph.ox.ac.uk/showcase/) in the UK. The UK Biobank proteomic assay was performed in the Pharma Proteomics Project (PPP) collaboratively funded by 13 biopharmaceutical companies, with a sample size of 54,219. ^20^ Among them, 53,014 had valid proteomic data release as of the time of analyses for the current study, and we excluded individuals aged <55 years at recruitment, with dementia at baseline or developed dementia within the first 2 years of follow-up, or with stroke at baseline. The participant inclusion flowchart was shown in **Supplementary Figure 1**.

The Framingham Heart Study (FHS) began in 1948 as one of the earliest population-based cohort studies.^43^ In 1971, the FHS initiated an offspring cohort by including 5,124 individuals consisting of offspring of the original cohort and their spouses. The FOS conducted proteomic assays using blood sample collected at Exam 5 in 1991-1995 (N=1856). We used similar inclusion/exclusion criteria as in UK Biobank (**Supplementary Figure 1**).

All participants provided written consent. UK Biobank was approved by the North West Multi-centre Research Ethics Committee (16/NW/0274). FHS was approved by the Boston Medical Center.

### Definitions of Healthy Lifestyles and the Healthy Lifestyle Score

We included seven healthy lifestyles according to existing literature for dementia prevention and followed existing literature in defining them in the UK Biobank.^11,44,45^ We defined no current smoking according to self-reports. Regular physical activity was defined as more than 150 minutes/week of moderate activity, 75 minutes/week vigorous activity, an equivalent combination, 5 days/week of moderate activity, or 1 day/week vigorous activity. A healthy diet is defined as meeting >=4/7 recommendations on fruit, vegetables, fish, whole grains, refined grains, red meats, and processed meats. Moderate alcohol consumption levels were >0-1 drinks for women and >0-2 drinks for men. Healthy sleeping duration was 7-8 hours/day. Low sedentary behaviour was defined as <=4 hours /d television watching, Adequate social activities was defined as attending social activities at least once per week. In the FHS offspring cohort, we slightly modified the criteria of physical activity (dichotomized by median of physical activity index^46^) and healthy diet (dichotomized by median of Dietary Guidelines for Americans Adherence Index, DGAI^47^). A weighted healthy lifestyle score was calculated based on regression coefficients of each component in the Cox proportional hazards model that included all seven lifestyle factors and adjusted for age at recruitment, sex, ethnicity, Townsend deprivation index, and highest education level, with a higher score indicating healthier lifestyle.

### Ascertainment of Dementia

UK Biobank used electronic health records with linkage to National Health Service to collect incident dementia and dementia-related mortality information was extracted from the death registry. The ascertainment algorithm was described in a previous study.^48^ Briefly, the International Classification of Diseases (ICD) and Read codes for all-cause and cause-specific dementia was identified from the electronic health records and death registry were combined to accurately define incident dementia. We also included the code definitions in **Supplementary Table 2**. Due to limited number of dementia cases in the FOS, we did not include the FOS dementia outcome.

### Proteomic Assays

UKB-PPP used Olink Explore 3072 antibody-based proximity extension assay. Details of participant selection and sample handling for this specific project has been described in a previous study.^20^ The protein level was inverse rank normalized and used in analysis as the normalized protein expression (NPX) value. Of the 2,923 proteins that were available in the latest data release, we excluded proteins with a missing value rate over 20% (i.e., detection rate ≥80%).^20^

At Exam 5, FOS collected plasma samples in citrate-treated tubes, centrifuged them 2000g for 10 min, aliquoted plasma samples and stored them at −80 °C. The samples were quantified for 1373 proteomic features using the aptamer-based SOMAScan v1.3 platform with single-stranded DNA-based aptamers in two batches.^49^ We analysed the proteins with further adjustment for batch effect, We mapped the proteins across the OLink and the SOMAScan platforms using Uniport ID.

### Covariates

The covariates of the current study included age (calculated from date of recruitment and date of birth), sex (self-identified), race or ethnicity (white or non-white, only available in the UK Biobank), highest educational level (categorized as college or above/high school of below), and socioeconomic status measured by Townsend deprivation index^50^ in UK Biobank or household income in the FOS. All covariates were collected concurrently with healthy lifestyle measurements. Health conditions included body mass index (BMI, calculated from objectively measured weight and height in physical examination), the Patient Health Questionnaire (PHQ)-2 score, diabetes, hypertension, cardiovascular disease (including myocardial infarction, stroke, and heart failure), and dyslipidaemia. All chronic diseases were ascertained through self-reports, physical examinations, blood biochemical assays and linkage to the electronic health records. We did not consider health conditions as primary covariates because they may be important mediating variables but included them in the model for sensitivity analyses.

### Statistical Analysis

Baseline characteristics of the sample population were summarized according to lifestyle score quintiles as percentages for categorical variables and as means and standard deviations for normally distributed continuous variables. Multiple imputations by chained equations with 25 imputations were used to impute missing values given the low missing rates (<5%) of the phenotypic variables.^51^

The primary analyses were implemented in the UK Biobank. The associations of the healthy lifestyle score and individual healthy lifestyle factors with each plasma protein were assessed using multivariable-adjusted linear regression models, with adjustment for the full covariate set. Cox proportional hazard models were used to assess the association of the healthy lifestyle score, individual lifestyle factors, and plasma proteins with future risk of dementia. The full set of covariate adjustments included age at recruitment, sex, race or ethnicity, Townsend deprivation index, and highest education level. Person-year at risk was calculated from the date of recruitment (2006-2010) to date of first diagnosis, death of other causes, loss to follow-up, or the end of follow-up, whichever came first. The Kaplan– Meier curves plots the cumulative probability of survival in a given period as a function of time. In the sensitivity analysis, we further adjusted the models for BMI, PHQ-2 score, prevalent diabetes, hypertension, cardiovascular disease, and dyslipidaemia. The proteins simultaneously significantly associated with healthy lifestyle score or healthy lifestyle factors and incident dementia with Bonferroni-corrected P-values were considered potential mediators for the lifestyle-dementia association. The mediation analysis was conducted within a causal framework by assessing the reduction proportion in coefficient of healthy lifestyle score or healthy lifestyle factors on dementia risk.^29^ We then adopted the Preacher and Hayes bootstrapping method^52^ (1000 times) to calculate the corresponding confidence intervals for proportion of mediation in a non-parametric test. We performed Gene Ontology (GO) enrichment analyses and Kyoto Encyclopedia of Genes and Genomes (KEGG) to investigate the potential pathways. To explore the potential interactions between identified proteins, a PPI network was constructed using the STRING database (https://string-db.org/). Further, we externally validated the associations of the above mediator proteins with lifestyle scores using the FOS cohort. As mentioned above, we did not include dementia outcomes in the FOS given the limited number of dementia cases, large-scale association analyses may have been underpowered.

We assessed the interactions of the protein signatures and healthy lifestyle score or healthy lifestyle factors in associations with incident dementia using the Cox proportional hazard models, with the same adjustment set as above. We determined potential protein modifiers by assessing the statistical significance of the crossing product term of the protein and the healthy lifestyle score or healthy lifestyle factors. We also accounted for multiplicity using Bonferroni correction as in the association analyses, but upscaled the threshold to Bonferroni-corrected P-values as 0.10 considering reduced power to detect interactions^21–23^.

To develop the LPSSs for the healthy lifestyle score and individual healthy lifestyle factors, we first identified proteins that showing nominal significant interactions with healthy lifestyle score or healthy lifestyle factors in the associations with dementia. This would leave 200-400 candidate proteins for LPSSs construction. We applied forward stepwise approach to select proteins starting with the most significant protein.^53^ We entered each protein and its interaction with lifestyle score or lifestyle factor into the regression model one by one. After each new protein was added, proteins that still interacted significantly in the model were retained, while proteins that no longer interacted significantly were eliminated. The LPSSs were calculated from the weighting coefficients of the interaction terms with healthy lifestyle score or factors in the final model. We randomly splited the study population into training and testing sets (1:1) for LPSSs development and strafication framework utility assessments. The ratio of 1:1 for splitting was selected to ensure a sufficient power for dementia analysis in the testing set. The weighting coefficients for features selected during train population were used to project scores into the test population. We further estimated the PAF from the estimated HR for the exposure and the proportion of persons in the population exposed to the risk factor (Pe), as PAF = Pe(HR – 1)/(1 + Pe[HR – 1]). All analyses and modelling were implemented using R (4.3.0). The pseudo code of this algorithm was described in **Supplementary Methods**.

---

**Supplementary Figure 1.**
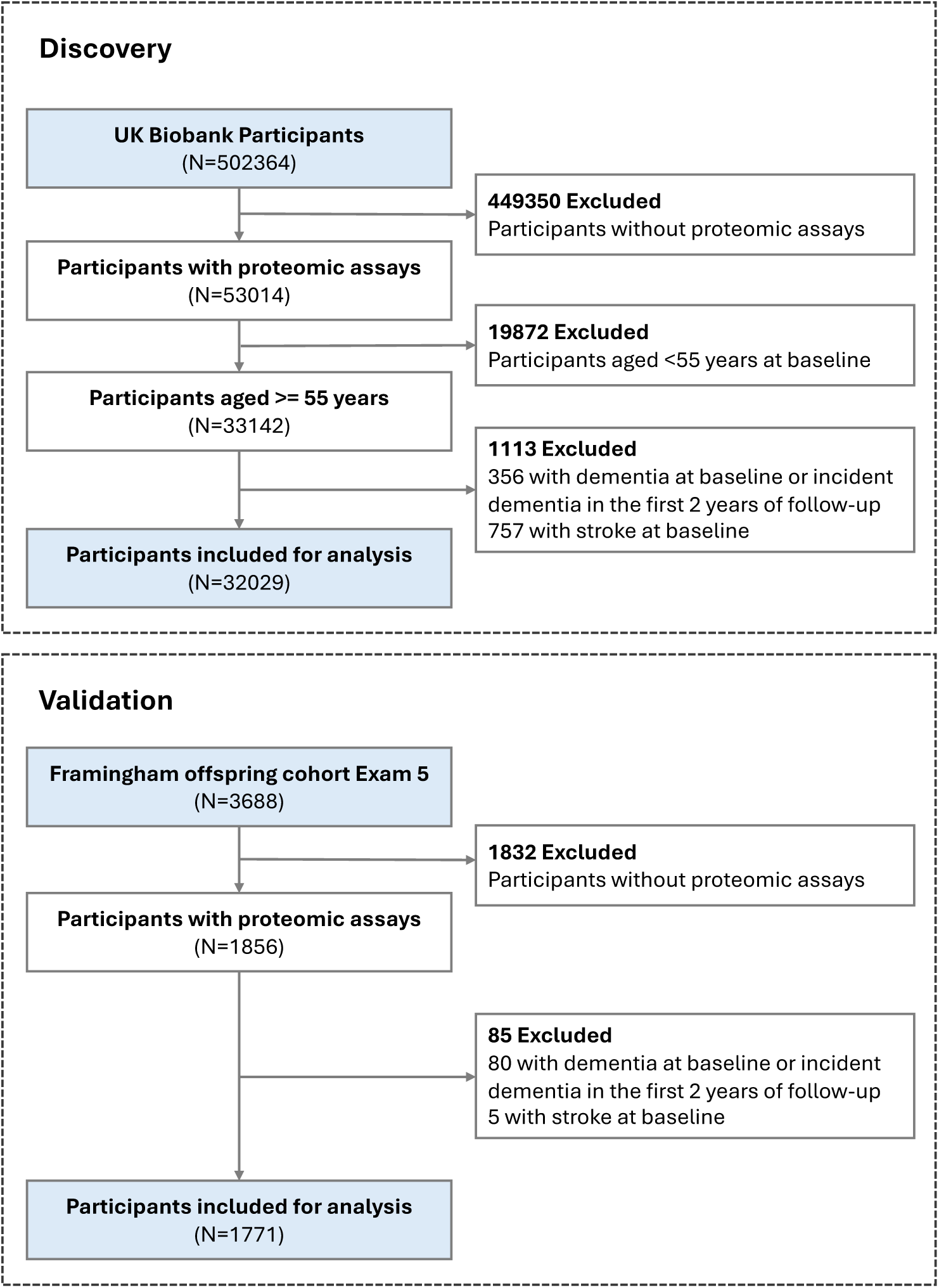
Flowcharts of participants inclusion in the current study

**Supplementary Figure 2.**
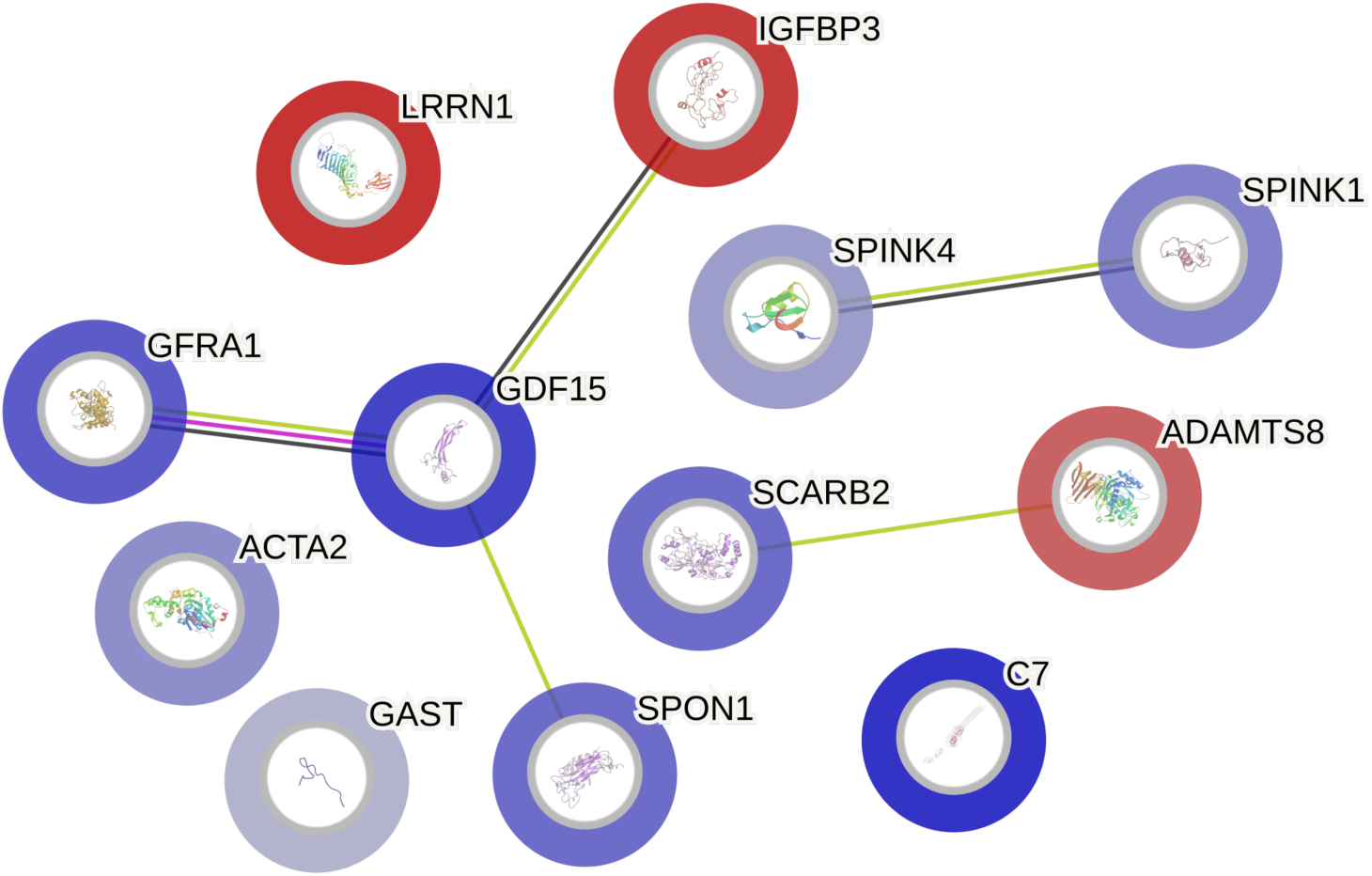
Protein–protein interaction network for 12 proteins that significantly associated with both the overall healthy lifestyle score and dementia risk Network nodes represent proteins. Edges represent protein-protein associations (Purple: known interactions with experimentally determined; Blue: predicted interactions with gene co-occurrence; Green: interactions with textmining; black: interactions with co-expression).

## Supplementary Methods

The pseudo code of this algorithm is as follows:

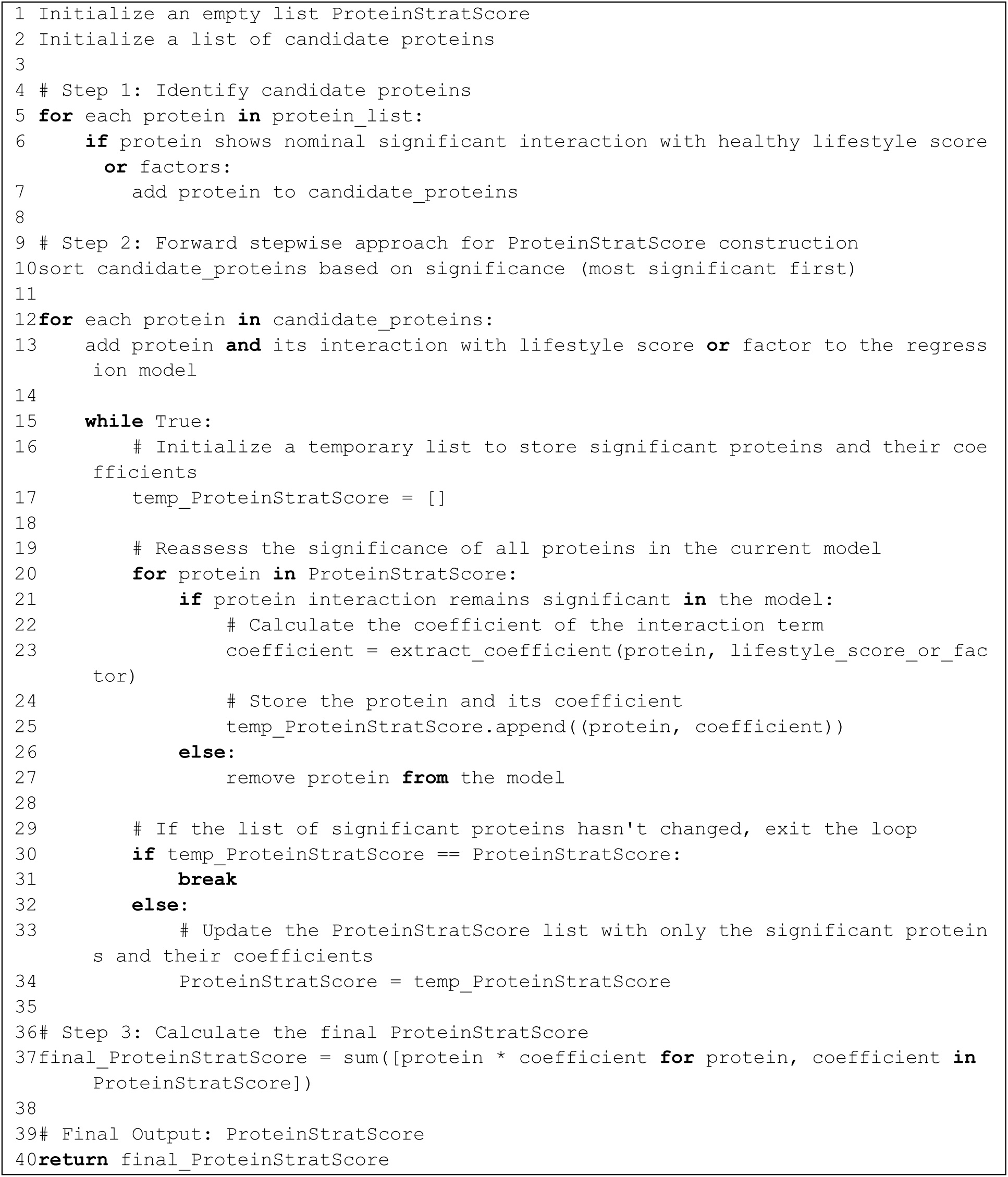

